# Drink more, sleep less: Applying compositional data analysis to investigate day-to-day associations between alcohol use and 24-hour movement behaviors

**DOI:** 10.64898/2026.07.28.26359097

**Authors:** Jimikaye B. Courtney, Georgiy V. Bobashev

## Abstract

**Background:** Alcohol use impacts sleep; however, sleep is also impacted by other 24-hour movement behaviors, including sedentary behavior (SB) and physical activity (PA). This study used compositional data analysis (CoDA) to simultaneously account for SB and PA and investigate within- and between-person associations between sleep and alcohol use behaviors.

**Methods:** Participants were 21-44-year-old overweight/obese adults. Across 21 days, participants wore an activPAL monitor to assess sleep, PA, and SB hours, and completed morning surveys of past day alcohol use to assess drinks/day. CoDA identified the daily proportions of time spent in sleep, PA, and SB. Multilevel models investigated within- and between-person associations between sleep, demographic factors, and alcohol use. Differences in sleep, SB, and PA across drinking versus non-drinking days were investigated with individual MANOVAs for each participant.

**Results:** Participants (N=91, M_age_=30.7±6.5, 57% female, 75% White) reported drinking on 23% (n=437) of days, consumed 1 drink/day, and averaged 8.5±2.3 sleep hours, 7.4±2.8 SB hours, and 8.0±2.7 PA hours per day. Multilevel models indicated that participants slept 16.6 fewer minutes on drinking days (*p*=.046) each additional drink corresponded with 4.4 fewer minutes of sleep (*p*=.04),. There was large between-person variability in time use allocations across drinking versus non-drinking days. For many the change was non-significant; however, three participants spent significantly less time sleeping on drinking days (1.16-4.74 fewer hours of sleep) and one participant spent significantly (2.62) more hours sleeping on drinking versus non-drinking days.

**Conclusions:** Drinking days and greater alcohol consumption were associated with fewer minutes of sleep within-people; however, shifts towards PA and SB were inconsistent. Investigating contextual factors, such as the physical/social context of alcohol use, the time-of-day drinking occurs, the type of alcohol consumed, or the type of PA a person engages in could provide deeper insight into the conditions under which alcohol use detrimentally impacts sleep.

## Introduction

The National Sleep Foundation recommends that adults 18-64 years old sleep 7 to 9 hours per night [1]. Unfortunately, 30-40% of adults in the United States, Australia, and European countries do not meet these sleep recommendations [2–4], and 10-28% of the adult population use alcohol as an over-the-counter sleep aid [5–7].

Alcohol acts as a sedative, with alcohol consumption exerting pronounced and clinically meaningful effects on sleep duration, architecture, and quality [8]. Acute alcohol consumption (i.e., alcohol consumed in the hours preceding sleep on a given day) decreases sleep onset latency, helping people fall asleep more quickly [8–11]. During the first half of the night, alcohol consumption also decreases wakefulness after sleep onset and increases the duration of non-rapid eye movement (REM) sleep [8–11]. However, during the second half of the night, alcohol consumption increases wakefulness after sleep onset, delays onset of REM sleep, and decreases the duration of REM sleep [9–11]. These acute effects of alcohol are typically dose-dependent, with heavier consumption yielding stronger effects [9–11]. Overall, acute alcohol consumption worsens sleep quantity and quality. From a chronic perspective, repeated alcohol use is associated with persistent insomnia symptoms, altered sleep architecture, and long-term reductions in restorative sleep stages (e.g., REM sleep) [8].

When considering the acute associations between alcohol use and sleep in daily life, it is important to consider that sleep does not occur in isolation. Rather, sleep is one component of 24-hour movement behaviors that also include sedentary behavior (SB) and physical activity (PA), both of which influence sleep [12–15]. PA improves sleep quality, sleep onset latency, total sleep time, sleep efficiency, wakefulness after sleep onset, and REM sleep [16,17]. The effects of PA on sleep are observed among both alcohol users and non-users [18]. SB has the opposite effect of PA on sleep, with greater SB increasing the risk of insomnia, disturbed sleep, and sleep problems [19,20]. Therefore, failure to consider PA and SB when investigating the associations between alcohol use and sleep could yield inaccurate findings or spurious correlations.

Because the components of the day necessarily sum to a finite total, 24-hour movement behaviors are completely co-dependent, such that increases in time spent in one behavior (e.g., sleep) imply decreases in time spent in other behaviors (e.g., SB or PA) [12–14]. This dependency complicates inference from conventional regression approaches when investigating how alcohol relates to sleep and other health outcomes [14,15]. Unfortunately, few existing studies investigating alcohol use associations with sleep account for SB or PA, violating the idea that the effects of time spent in one domain of 24-hour movement behaviors can only be fully understood when considered in terms of the functional relationships with time spent in other 24-hour movement behavior domains [13].

Compositional data analysis (CoDA) provides a statistically principled framework for modeling time-use data by representing behaviors as parts of a whole and using log-ratio transformations to capture relative time reallocations among sleep, SB, and PA [13]. Application of CoDA has clarified relationships between 24-hour movement behavior compositions and cardiometabolic and other health endpoints and is well suited to studies examining how alcohol use co-varies with the distribution of daily 24-hour movement behaviors [21,22].

Wearable sensors have transformed the measurement of 24-hour movement behaviors under free-living conditions by enabling continuous, objective, and high-resolution assessment over days to weeks. Compared with polysomnography in laboratory settings, many consumer and research-grade wearables demonstrate acceptable agreement for sleep/wake classification and estimates of sleep duration in population studies, while providing reliable measurement of SB and various intensities of PA [23–28]. Combining wearable sensors with intensive longitudinal methods (i.e., studies that include frequent and dense repeated measures captured across acute timescales, such as every second, hour, or day [29,30]) to assess daily drinking behaviors enables the evaluation of temporal relationships between drinking episodes and subsequent sleep alterations in real-world settings. These capabilities facilitate within-person (day-level) analyses of how drinking on a given day pertains to the following night’s sleep while simultaneously accounting for the composition of the remaining 24-hour movement behaviors (i.e., SB, PA).

Previous studies have investigated whether daily alcohol use predicts nighttime sleep [31–37]. Tussey et al., (2025) found no within-person associations between daily drinking and nighttime sleep; however, greater average levels of drinking were associated with feeling less rested, greater average sleep time, and greater wakefulness after sleep onset. Other studies did find within-person effects, such that greater than usual daily alcohol consumption was associated with worse nighttime sleep quality, delayed bed and wake times, and greater wakefulness after sleep onset [31–34,36]. Additionally, on days individuals engaged in binge drinking they reported less nighttime sleep, worse sleep quality, and greater next day tiredness [35]. Unfortunately, none of these studies accounted for other 24-hour movement behaviors (i.e., SB, PA), which is problematic given the effects of SB and PA on sleep [16–20], as well as the co-dependency between sleep, SB, and PA [12–14].

Together, these factors motivated our investigation to use a recent pilot study to investigate the effect sizes for a hypothesis that alcohol would result in greater overall sleep duration, with greater sleep on drinking days, and a dose-dependent association between alcohol and sleep. We aimed to (1) quantify how alcohol consumption relates to same day sleep using objective, continuous measures in daily life and (2) frame those associations within the full 24-hour composition of sleep, SB, and PA using CoDA methods. Such an approach can help identify intervention targets to ameliorate alcohol-related sleep disruption and its downstream health consequences.

## Materials and Methods

### Participants, recruitment, and screening

Participants were drawn from a larger study that specifically recruited 91 adults aged 21–44 years with overweight or obesity over a 19-month period (April 2023–October 2024) [38]; the present manuscript represents secondary analyses of those data. Participants were recruited from the southeastern U.S. via campus flyers, class announcements, emails, and social media. Eligibility criteria included being 21–44 years old, English-speaking, overweight/obese (body mass index [BMI] ≥ 25 kg/m² based on self-reported height and weight), and owning an iPhone 6S or newer. Participants were classified into drinker (≥ 1 alcoholic beverage/week) and non-drinker (0 alcoholic beverages/week) cohorts. Exclusion criteria included inability to perform unassisted PA, competitive athletics, pregnancy or breastfeeding, history of alcohol use disorder, or diagnosis of cancer, cardiovascular disease, or diabetes. Of 662 individuals screened via Research Electronic Data Capture (REDCap) [39], 231 (34.9%) qualified, 94 (14.2%) consented, and 91 (96.8%) completed the study. The study was approved by the Institutional Review Board at the University of North Carolina at Chapel Hill, and participants provided written informed consent.

### Procedures

Participants completed a baseline visit with surveys, anthropometric measurements, and training on how to use study devices, followed by a 21-day field protocol that included device wear (activPAL physical activity monitor, BACtrack Skyn alcohol monitor) and daily morning reports of past day alcohol consumption, and a follow-up visit with surveys and a blood draw. More details of the study protocol and measures can be found elsewhere [40].

### Measures

#### Demographics

Participants self-reported demographics via an electronic baseline survey, including biological sex, race, ethnicity, student status, education, employment, and income [41].

#### Anthropometrics

Participant anthropometrics (height [cm], weight [kg], waist and hip circumference [cm]) were measured in duplicate with heavy clothing and shoes removed. Body mass index (BMI) was calculated as weight (kg) ÷ height (m)^2^.

#### Alcohol use

Habitual alcohol use was reported during the screening survey via the Daily Drinking Questionnaire [42], a widely used measure of alcohol use with demonstrated construct validity including consistent associations with alcohol-related consequences and other indices of drinking behavior [43,44]. Participants were categorized as non-drinkers (n=29, 31.9%) if they reported consuming no alcohol over the past month. Participants were categorized as light drinkers if they consumed ≤3 drinks/week (n=17, 18.7%), moderate drinkers if they consumed 4-7 (females) or 4-14 (males) drinks/week (n=23, 25.3%), or heavy drinkers if they consumed 8+ (females) or 15+ (males) drinks/week (n=22, 24.2%). Daily alcohol use was assessed via daily smartphone surveys sent each morning at 9AM. Participants reported the amount and type of alcohol consumed the previous day, with standard drink servings defined as 12 ounces (oz) of 5% alcohol by volume beer, 8-9 oz of 7% beer, 4-5 oz of wine, a 1.5 oz shot of hard liquor, or 12 oz of hard seltzer, and a picture showing the different drink sizes. The total number of drinks consumed per day was calculated from the self-reports, and we lagged values one day to ensure that they aligned with the same lived day as the 24-hour movement data. A day was defined as a ‘drinking day’ if a participant reported consuming one or more alcoholic beverages. If they reported consuming no alcoholic beverages, it was defined as a ‘non-drinking day’. Participants demonstrated high compliance, completing surveys on 89% of study days (n=1,699).

#### 24-hour movement behaviors

24-hour movement behaviors were measured using the activPAL accelerometer. The activPAL is a validated triaxial accelerometer and inclinometer that measures step counts, postural position, and acceleration in 20 second epochs [45,46]. The activPAL was water-proofed with a nitrile sleeve and worn on the non-dominant thigh for 21 consecutive days, with participants only removing the device to bathe or swim. activPAL data were processed using the CREA algorithm to assess daily time spent in sleep, moderate to vigorous physical activity (MVPA) and light physical activity (LPA), and sedentary behavior (SB). Rather than assessing daily values using a traditional calendar day (12:00AM-11:59PM), which would have resulted in some of the sleep occurring prior to alcohol use, we defined a ‘study day’ as 9:00AM on day *n* to 8:59 AM on day *n*+1 to align with the daily alcohol use reports and ensure that alcohol use preceded sleep.

A wear day was considered valid if the participant wore the activPAL for at least 20 hours of total wear time, including a minimum of 10 waking hours and non-zero sleep duration (n=1655 days, 83.4%). Participants needed to have three valid weekdays and one valid weekend day to be included in the analyses. Five of the 91 participants did not meet these criteria, resulting in a final analytic sample included 86 participants. Among participants meeting wear-time criteria, an average of 19.8 ± 2.2 valid days were recorded per participant. Mean daily wear time was 23.98 ± 0.26 hours (range: 20.01-24.00 hours), indicating that most days reflected near-complete 24-hour monitoring (see Supplemental Table 1 for full details on device wear). The CREA algorithm was used to derive posture- and movement-based classifications [47,48]. The algorithm’s primary lying time variable was used to estimate nighttime sleep duration, consistent with standard use as a proxy for bed/sleep time [47]. SB while awake was defined using the CREA-derived sedentary estimate, which includes time spent sitting, in seated transport (e.g., driving) and secondary lying time. Secondary lying time was included in SB because it reflects waking behaviors such as reclining or daytime napping, and is incorporated within activPAL’s default SB classification. PA was classified as MVPA when stepping cadence was ≥100 steps per minute [49,50], with total daily MVPA duration calculated accordingly. To ensure internal consistency, such that all behavioral components summed to a full day, LPA was derived as the residual time within the observed day (i.e., LPA = total daily wear time – [sleep + SB + MVPA]). Non-wear time was identified using the CREA algorithm, which detects sustained periods of minimal acceleration and the absence of postural transitions to identify non-wear [48]. Importantly, total daily wear time reflects observed accelerometer wear time rather than a standardized 24-hour day, and non-wear time was not imputed. Thus, movement behavior compositions were based on observed wear-time data and were not normalized to 24 hours. Given our focus on sleep outcomes, MVPA and LPA were combined into a single physical activity component to reduce compositional complexity and facilitate interpretation of relative time-use patterns so that the models included three components of 24-hour movement behaviors: sleep, SB, and PA. The 24-hour movement behaviors from each valid wear day were used in the analyses.

### Statistical analysis

We used compositional data analysis (CoDA) to characterize and model the daily distribution of time spent in sleep, PA, and SB. Because time-use data are constrained to a 24-hour period, with each component representing a proportion of the fixed 24-hour period, standard statistical methods assuming independence among variables are inappropriate [13]. To address this, daily durations of sleep, PA, and SB were converted to proportions of total daily time (e.g., 8 hours of sleep = 33.3% of 24-hour day) [51]. While we considered both isometric log-ratio (ILR) transformations and additive log-ratio (ALR) transformations for consistency of the analysis we used ILRs transformations for both, the multi-level models and the multivariate analysis of variance models. ILRs express movement behaviors as independent orthogonal coordinates while preserving their relative structure for valid use in linear regression models [52]. The ILR coordinates included: ILR1) sleep versus SB + PA, and ILR2) SB versus PA. The ILR2 coordinate for SB versus PA was used as a covariate in the multi-level models. Additionally, ILR transformation is consistent with Atchison distances in ternary plots. Both ILR coordinates were used as dependent variables in the multivariate analysis of variance models.

We applied multilevel linear regression models to examine both within-person (day-level) and between-person (individual-level) associations between alcohol use and sleep time, accounting for SB and PA (ILR2 coordinate), and demographic factors. The hierarchical structure nested daily observations within individuals (i.e., random intercepts) to account for the repeated measures. Because drinking days were also nested within an individual, random slopes were included to investigate interindividual variability in drinking associations with sleep. The within-person component of the model estimated associations between day-to-day variations in daily alcohol consumption and subsequent nighttime sleep, with day-level values centered to participant means. The between-person component of the model estimated associations between average alcohol consumption and average sleep time, with participant mean values centered to the sample mean. Demographic characteristics (age, sex, race) as well as weekend (Saturday, Sunday) versus weekday (Monday-Friday) were also tested. Models were estimated using restricted maximum likelihood (REML) with robust standard errors. Model diagnostics included evaluation of residual distributions, homoscedasticity, and intraclass correlation coefficients (ICCs) to quantify the proportion of variance attributable to between-person differences. We conducted models with the entire sample, as well as sensitivity analyses in the drinking cohort. To evaluate potential nonlinear associations, we compared spline regression models with 1 to 3 degrees of freedom for both within-person and between-person predictors, using likelihood ratio tests and information criteria to assess model fit. These comparisons indicated no meaningful improvements in fit with increased flexibility; therefore, linear models were used for all subsequent analyses. Although CoDA was used to derive ILR coordinates, sleep time (hours) was modeled as an absolute outcome to address the research question focused on sleep time. Including ILR2 as a covariate partially accounts for the relative distribution of non-sleep behaviors, but the regression model results should be interpreted as associations with absolute sleep time rather than compositional reallocations. All multi-level models were conducted using the lme4 package in R.

We conducted exploratory heterogeneity analysis by using individual multivariate analysis of variance (MANOVA) models on 51 participants in the drinking cohort who had at least six days of data with at least one drinking day during the study period and who wore the activPAL for at least 23 hours per day. For each individual participant the MANOVAs compared time use compositions of 24-hour movement behaviors across the participant’s drinking and non-drinking days using the ILR coordinates. For participants with significant MANOVA findings, follow-up one-way ANOVAs compared the relative proportions of sleep versus SB + PA (ILR1) and sleeps versus PA (ILR2) on drinking versus non-drinking days. We created ternary plots displaying each participant’s daily observations as a point inside an equilateral triangle with the top vertex representing 100% PA time, the bottom left vertex representing 100% sleep time, and the bottom right vertex representing 100% SB time. All MANOVAs were conducted using R and statistical significance was set at *p* < .05

## Results

The summary of sample is Participant demographics are presented in Table 1. The mean ± standard deviation (SD) age of the participants (N=91) was 30.7 ± 6.52 years, and 57.1% were female (n=52), 74.7% were White (n=68), and 93.4% were non-Hispanic (n=85). Approximately one-third of participants were students (n=28), employed part-time (n=30), and had an income ≥$75,000/year (n=34). Most participants were either overweight (n=57, 62.6%) or obese (n=32, 35.1%) based on measured body mass index.

**Table 1.** Participant demographics.

| Characteristic | Non-Drinkers | Drinkers | All Participants |
| --- | --- | --- | --- |

|  | (N=29) | (N=62) | (N=91) |
| --- | --- | --- | --- |
| Age in Years (Mean ± SD) | 35.02 ± 5.94* | 28.69 ± 5.78 | 30.71 ± 6.52 |
| Sex (n (%)) |  |  |  |
| Male | 13 (44.8%) | 26 (41.9%) | 39 (42.9%) |
| Female | 16 (55.2%) | 36 (58.1%) | 52 (57.1%) |
| Race (n (%))<sup>a</sup> |  |  |  |
| White | 19 (65.5%) | 49 (79.0%) | 68 (74.7%) |
| Black | 5 (17.2%) | 5 (8.1%) | 10 (11.0%) |
| Asian | 2 (6.9%) | 6 (9.7%) | 8 (8.8%) |
| Mixed Race<sup>b</sup> | 1 (3.4%) | 1 (1.6%) | 2 (2.2%) |
| Ethnicity (n (%))<sup>c</sup> |  |  |  |
| Non-Hispanic | 27 (93.1%) | 58 (93.5%) | 85 (93.4%) |
| Hispanic | 1 (3.4%) | 4 (6.5%) | 5 (5.5%) |
| Student Type (n (%)) |  |  |  |
| Undergraduate | 3 (6.9%) | 11 (17.7%) | 13 (14.3%) |
| Graduate<sup>d</sup> | 2 (6.9%) | 13 (21.0%) | 15 (16.5%) |
| Non-Student | 25 (86.2%) | 38 (61.3%) | 63 (69.2%) |
| Employment Status (n (%))<sup>e</sup> |  |  |  |
| Full-Time | 16 (55.2%) | 31 (50.0%) | 47 (51.6%) |
| Part-Time | 6 (20.7%) | 24 (38.7%) | 30 (33.0%) |
| Unemployed | 7 (24.1%) | 7 (11.3%) | 14 (15.4%) |
| Income Status (n (%))<sup>f</sup> |  |  |  |
| ≤\$27,000 | 4 (13.8%) | 18 (29.0%) | 22 (24.2%) |
| \$27,000-\$43,999 | 3 (10.3%) | 6 (9.7%) | 9 (9.9%) |
| \$44,000-\$59,999 | 3 (10.3%) | 8 (12.9%) | 11 (12.1%) |
| \$60,000-\$74,999 | 6 (20.7%) | 5 (8.1%) | 11 (12.1%) |
| ≥\$75,000 | 13 (44.8%) | 21 (33.9%) | 34 (37.4%) |
| Body Mass Index in kg/m<sup>2</sup> (BMI)<sup>g</sup> |  |  |  |
| BMI (Mean ± SD) | 33.06 ± 8.85* | 29.45 ± 4.59 | 30.60 ± 6.44 |
| BMI Category (n (%)) |  |  |  |
| Normal Weight | 0 (0.0%) | 2 (3.2%) | 2 (2.2%) |
| Overweight | 16 (55.2%) | 41 (66.1%) | 57 (62.6%) |
| Obese | 13 (44.8%) | 19 (30.6%) | 32 (35.1%) |

Notes: \* $p < .05$ for difference between drinkers and non-drinkers. Standard Deviation, SD. <sup>a</sup> Missing race: n=3 (3.3%). <sup>b</sup> Mixed Race category includes participants who reported identifying as more than one race category. <sup>c</sup> Missing ethnicity: n=1 (1.1%). <sup>d</sup> Graduate student includes masters, doctoral, and professional students (e.g., law school student, medical school student). <sup>e</sup> Part-time includes both student and part-time employee. Unemployed includes unemployed non-students and students not working. <sup>f</sup> Income status is combined family income per year. Missing income: n=4 (4.4%). <sup>g</sup> Two participants who self-reported a BMI $\geq 25$ kg/m<sup>2</sup> in the screening survey had measured BMIs of 24.67 and 24.76 kg/m<sup>2</sup>, and they were allowed to remain in the study.

Table 2 shows participant descriptive statistics for alcohol use and 24-hour movement behaviors. The geometric mean composition of time use in the full sample was 8.90 hours of sleep, 7.17 hours of SB, and 7.91 hours of PA. Supplemental Table 2 presents the variation matrix (log-ratio variance) for movement behaviors, indicating the relative variability and co-dependence among components (Supplemental Table 2).

**Table 2.**
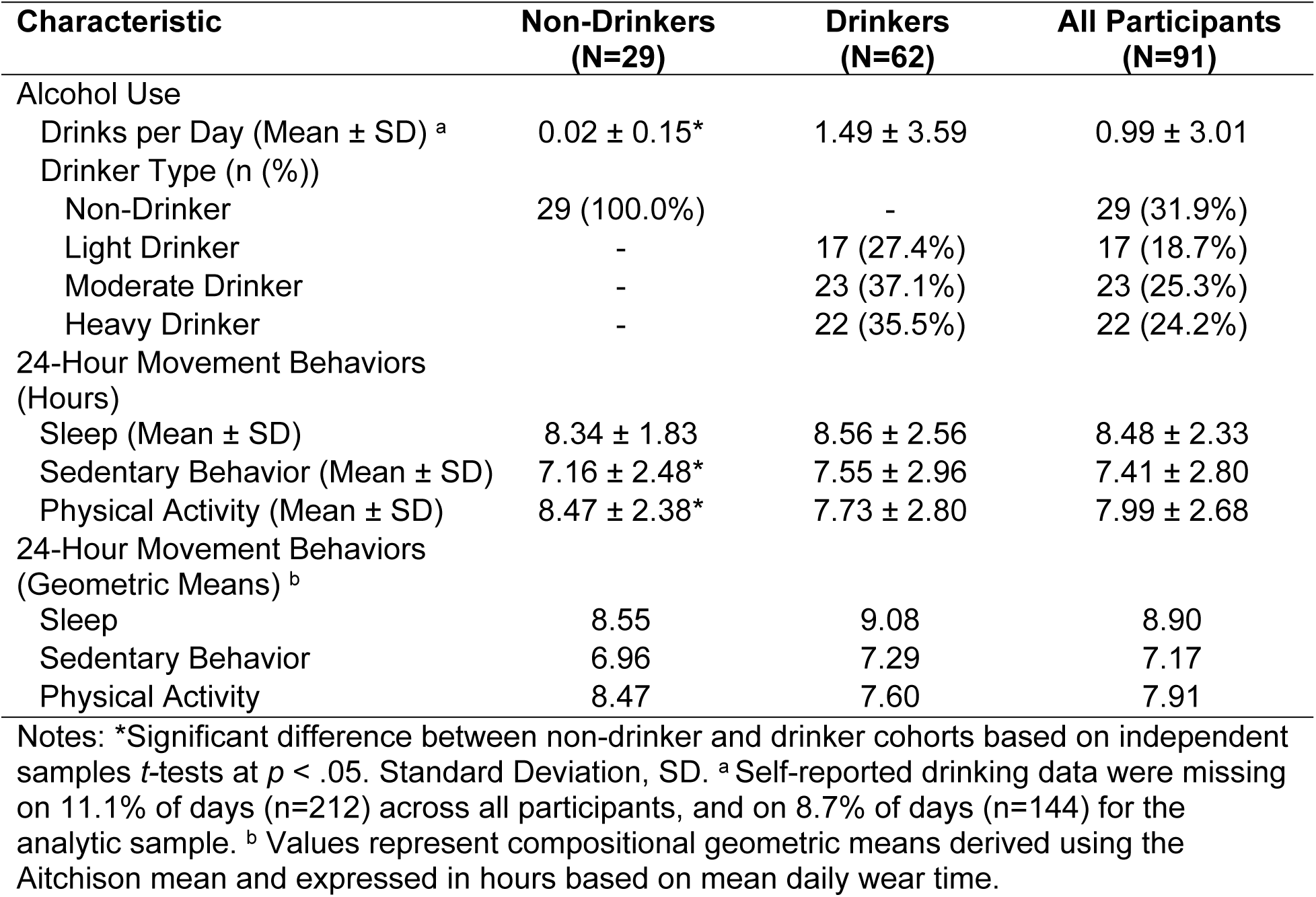
Participant descriptive statistics.

| Characteristic | Non-Drinkers<br>(N=29) | Drinkers<br>(N=62) | All Participants<br>(N=91) |
| --- | --- | --- | --- |
| Alcohol Use |  |  |  |
| Drinks per Day (Mean ± SD) <sup>a</sup> | 0.02 ± 0.15* | 1.49 ± 3.59 | 0.99 ± 3.01 |
| Drinker Type (n (%)) |  |  |  |
| Non-Drinker | 29 (100.0%) | - | 29 (31.9%) |
| Light Drinker | - | 17 (27.4%) | 17 (18.7%) |
| Moderate Drinker | - | 23 (37.1%) | 23 (25.3%) |
| Heavy Drinker | - | 22 (35.5%) | 22 (24.2%) |
| 24-Hour Movement Behaviors<br>(Hours) |  |  |  |
| Sleep (Mean ± SD) | 8.34 ± 1.83 | 8.56 ± 2.56 | 8.48 ± 2.33 |
| Sedentary Behavior (Mean ± SD) | 7.16 ± 2.48* | 7.55 ± 2.96 | 7.41 ± 2.80 |
| Physical Activity (Mean ± SD) | 8.47 ± 2.38* | 7.73 ± 2.80 | 7.99 ± 2.68 |
| 24-Hour Movement Behaviors<br>(Geometric Means) <sup>b</sup> |  |  |  |
| Sleep | 8.55 | 9.08 | 8.90 |
| Sedentary Behavior | 6.96 | 7.29 | 7.17 |
| Physical Activity | 8.47 | 7.60 | 7.91 |

Notes: \*Significant difference between non-drinker and drinker cohorts based on independent samples *t*-tests at $p < .05$ . Standard Deviation, SD. <sup>a</sup> Self-reported drinking data were missing on 11.1% of days (n=212) across all participants, and on 8.7% of days (n=144) for the analytic sample. <sup>b</sup> Values represent compositional geometric means derived using the Aitchison mean and expressed in hours based on mean daily wear time.

### Multi-level models for the association between alcohol use and sleep

Table 3 shows the models regressing sleep hours on drinking days and alcohol drinks/day. There was substantial within-person variability in daily sleep, with 74.1% of variance due to within-person differences (ICC=0.239). There was a statistically significant within-person effect of drinking days. Specifically, on days participants reported drinking alcohol, they slept for 16.6 fewer minutes (*b*=-0.28, *p*=.046). There was a statistically significant within-person effect of alcohol consumption such that, on days participants consumed one more alcoholic drink than their usual amount, they slept 4.4 fewer minutes (*b*=-0.07, *p*=.035). Participants who were older slept fewer hours (*b*=-0.05, *p*<.001), and participants slept 37.2 more minutes on weekends versus weekdays (*b*=0.62, *p*<.001). There were no statistically significant between-person associations between alcohol use and sleep time. The direction and significance pattern of the within-person alcohol coefficients were similar in the drinking-only cohort.

**Table 3.** Associations between daily and average alcohol use with sleep time (hours).

| **Models with All Participants** |  |  |
| --- | --- | --- |
|  | **Drinking Day <sup>d</sup>** | **Alcohol Drinks per Day <sup>d</sup>** |
|  | *b* (SE) | *b* (SE) |
| Intercept | 10.08 (0.47)*** | 10.04 (0.44)*** |
| Within-Person Effect on Sleep <sup>a</sup> | -0.28 (0.13)* | -0.07 (0.03)* |
| Between-Person Effect on Sleep <sup>b</sup> | 0.22 (0.36) | 0.04 (0.05) |
| Age | -0.05 (0.01)** | -0.05 (0.01)*** |
| Weekend | 0.60 (0.09)*** | 0.62 (0.09)*** |
| ILR2 (SB versus PA Time) <sup>c</sup> | -1.72 (0.11)*** | -1.70 (0.10)*** |

| **Sensitivity Analyses: Drinking Cohort** |  |  |
| --- | --- | --- |
|  | **Drinking Day <sup>e</sup>** | **Alcohol Drinks per Day <sup>e</sup>** |
|  | *b* (SE) | *b* (SE) |
| Intercept | 10.57 (0.69)*** | 10.43 (0.64)*** |
| Within-Person Effect on Sleep <sup>a</sup> | -0.25 (0.13)¥ | -0.05 (0.02)*** |
| Between-Person Effect on Sleep <sup>b</sup> | 0.61 (0.52) | 0.09 (0.08) |
| Age | -0.07 (0.02)** | -0.06 (0.02)** |
| Weekend | 0.61 (0.12)*** | 0.63 (0.12)*** |
| ILR2 (SB versus PA Time) <sup>c</sup> | -1.72 (0.13)*** | -1.72 (0.13)*** |

Notes: \*\*\* $p<.001$ ; \*\* $p<.01$ ; \* $p<.05$ ; ¥ $p<.10$ . Sedentary behavior, SB; Physical activity, PA; Unstandardized beta, *b*; Standard error, SE. <sup>a</sup> Within-person effects are day-level measures centered on participant means. For example, the day-level number of drinks consumed centered to the participant mean number of drinks consumed. <sup>b</sup> Between-person effects are participant means centered to the grand (sample) mean. For example, the participant mean number of drinks consumed per day centered to the sample mean. <sup>c</sup> This is the isometric log-transformed value for the relative proportion of time spent in sedentary behavior versus physical activity, which was included as a covariate to account for compositional structure. This was not interpreted independently. The other component of the balance between sleep versus sedentary behavior + physical activity time was not included due to perfect collinearity with the outcome – sleep hours. <sup>d</sup> Model included 1511 observations nested in 86 participants. There were random slopes due to improved model fit. There were no significant effects of sex, education, income, work status, race, ethnicity, or BMI. <sup>e</sup> Model included 949 observations nested in 57 participants. There was no random slope due to singularity issues. There were no significant effects of sex, education, income, work status, race, ethnicity, or BMI.

### MANOVAs comparing time-use compositions between drinking and non-drinking days

The MANOVA models were significant for five of the 51 participants (9.8%). Table 4 shows the results of the MANOVA models that were statistically significant. Follow-up ANOVAs were interpreted using reconstructed time-use differences to facilitate interpretation of compositional results. Although statistical tests were conducted on ILR-transformed variables, the original compositional parts were reconstructed using the inverse ILR transformation, and absolute differences in sleep, SB, and PA were expressed in hours to represent meaningful changes in daily time use. Follow-up analyses revealed heterogenous patterns across participants. Four participants showed reductions in sleep on drinking days compared to non-drinking days (ranging from 1.16 to 4.74 fewer hours) (Figs 1A-1D), although the difference was statistically significant only for participant 297. Specifically, participant 297 spent 4.74 fewer hours sleeping on drinking days, with time redistributed to SB (+0.86 hours) and PA (+3.88 hours) (Fig 1A). In contrast, participant 122 exhibited a shift in waking behavior, spending more time in PA (+4.26 hours of PA) and less time in SB (−3.10 hours) on drinking days, without a significant change in sleep (Fig 1B). Participant 147 slept significantly more hours on drinking days (+2.14 hours) compared to non-drinking days, spending less time in SB (−3.47 hours) and more time in PA (1.33 hours) (Fig 1E). Participants 37 and 271 showed differences in time use across drinking and non-drinking days (e.g., participant 271 slept 2.21 fewer hours on drinking days), but the follow-up analyses were not statistically significant (Figs 1C and 1D).

**Fig 1.**
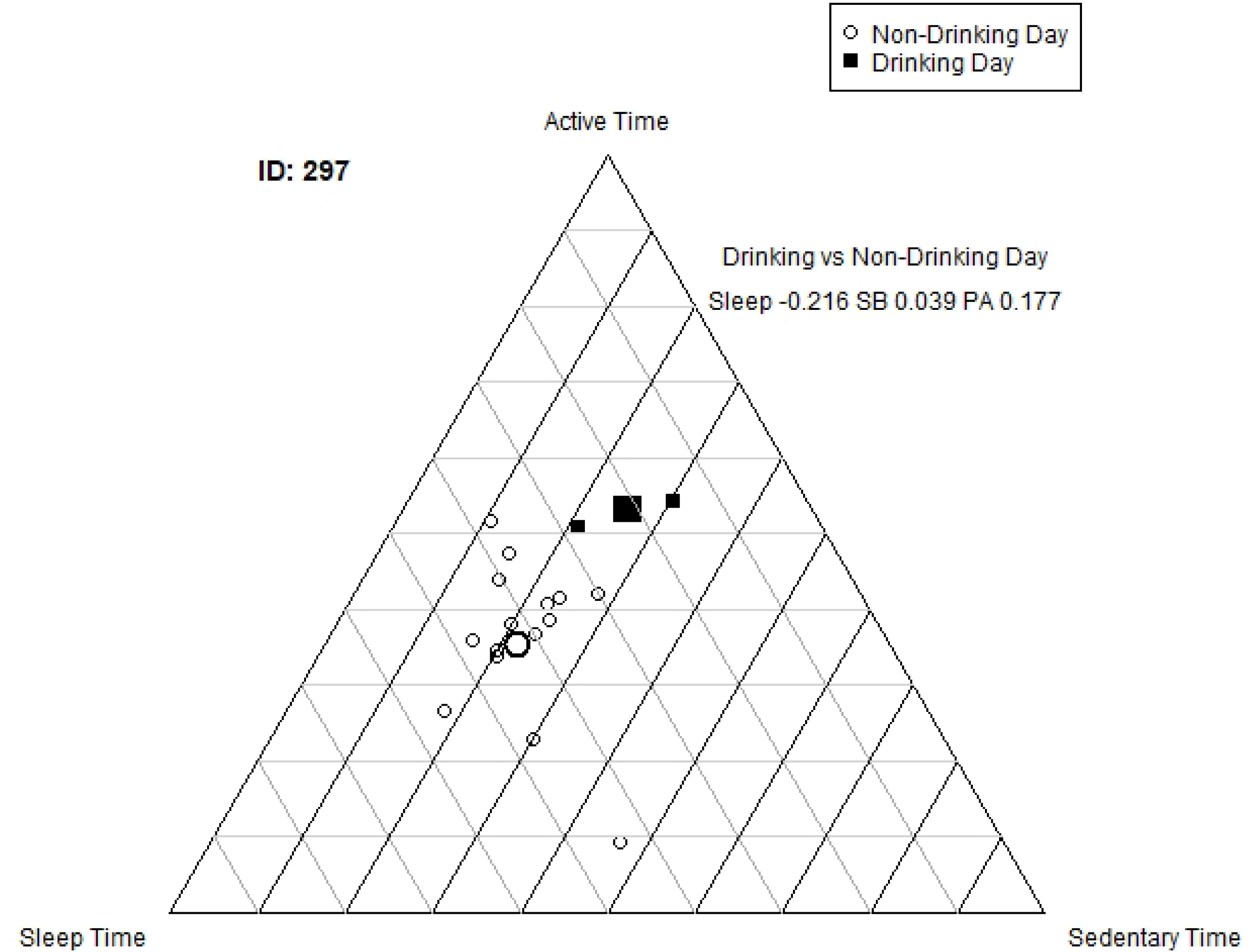
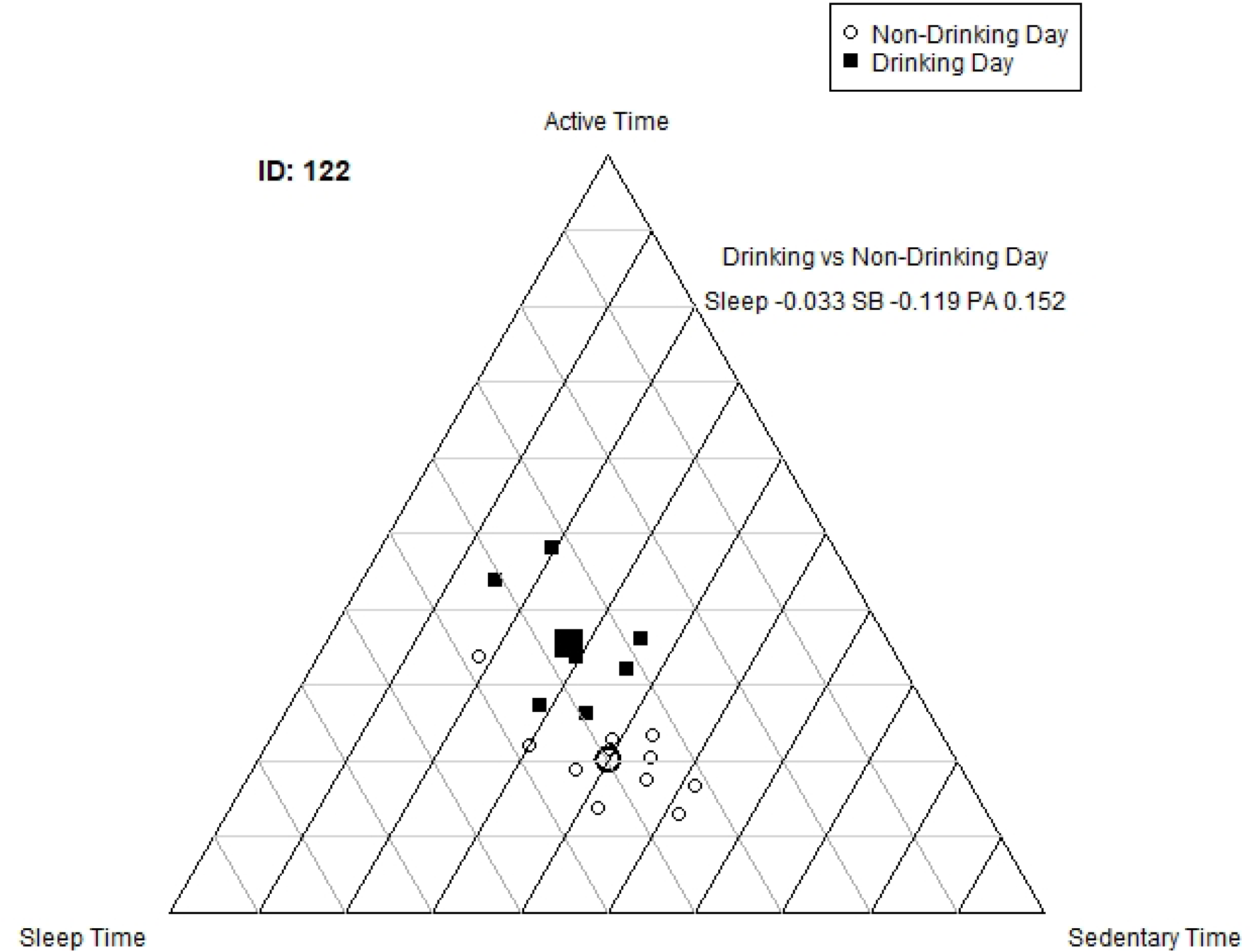
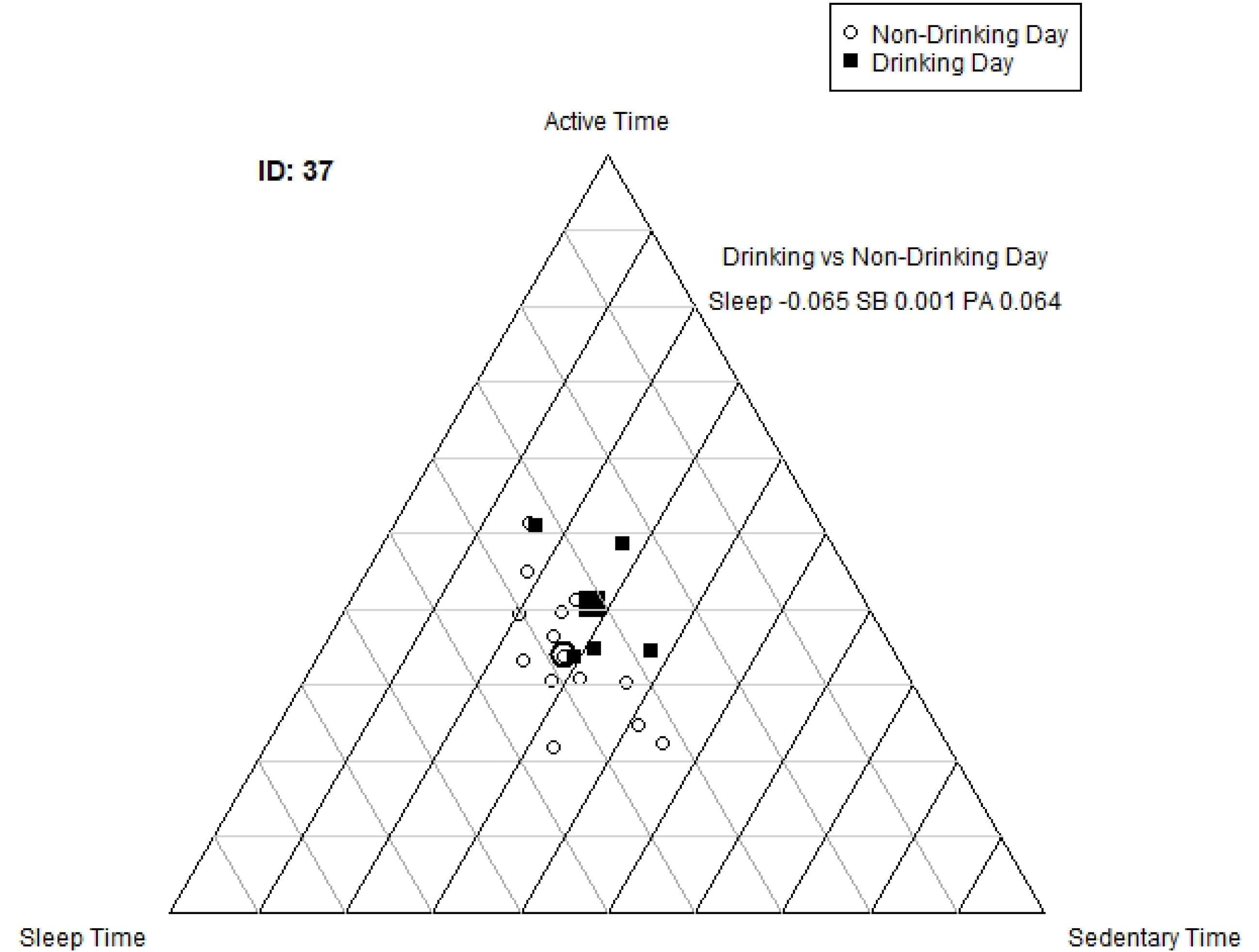
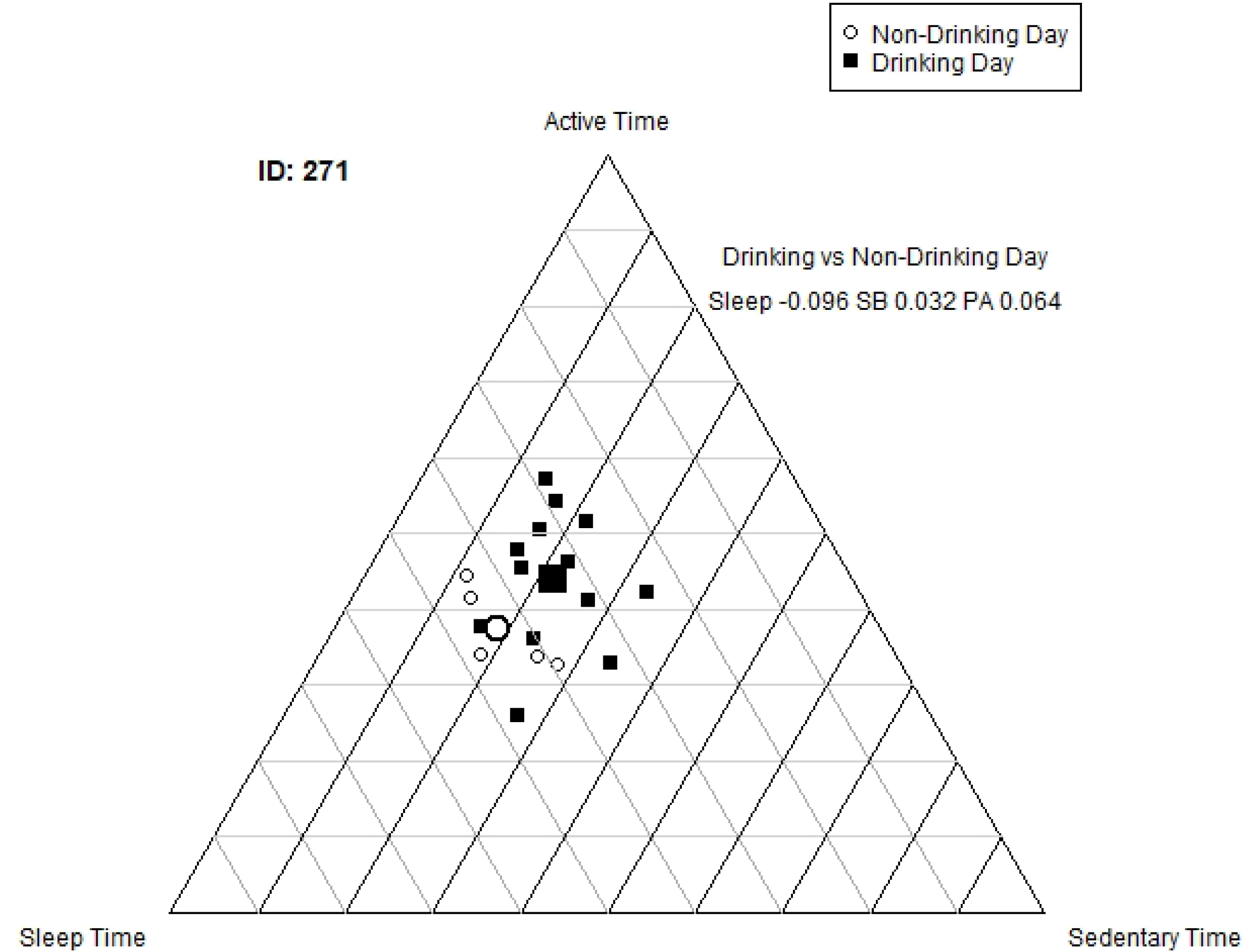
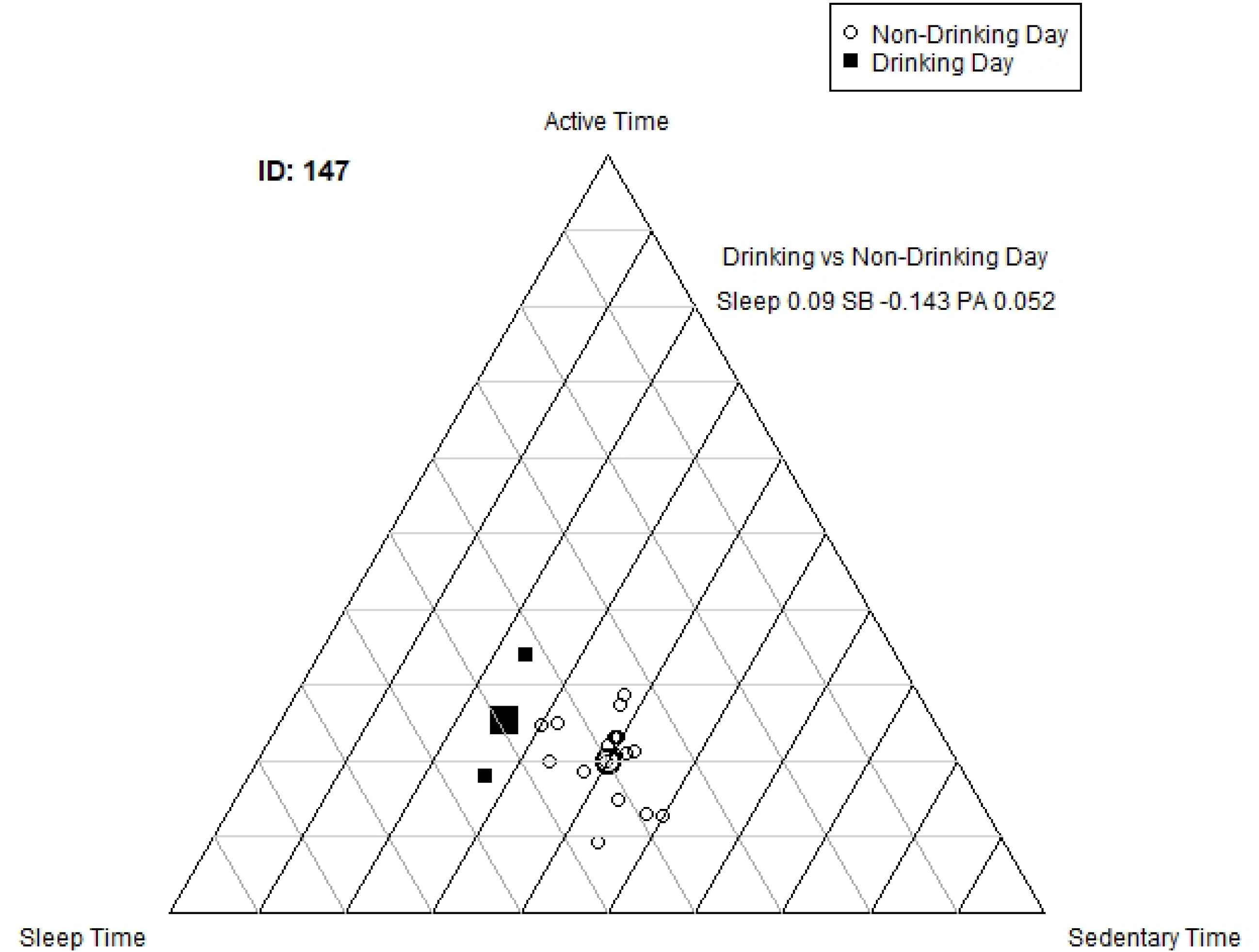
Ternary plots for 24-hour movement behaviors on drinking versus non-drinking days. 1A-1D. Ternary plots for participants 297 (1A), 122 (1B), 37 (1C) and 271 (1D) with decreased sleep during drinking days. Ternary plot for participant 147 with increased sleep during drinking days (1E).

**Table 4.** Individual differences in daily 24-hour movement behavior compositions between drinking and non-drinking days.

| MANOVA Results<br>for Drinking versus<br>non-Drinking Days <sup>a</sup> |  |  | Follow-Up One-Way ANOVA Results for<br>Drinking versus Non-Drinking Days <sup>a</sup> |  |  |  |  |
| --- | --- | --- | --- | --- | --- | --- | --- |
| ID | N Days | $F_{df1,df2}$ | ILR1 –Sleep<br>versus Wake<br>(SB+PA) (F) | ILR2 – SB versus<br>PA (F) | ΔSleep<br>(hours) | ΔSB<br>(hours) | ΔPA<br>(hours) |
| 37 | 19 | 4.17<sub>2,16</sub>* | 1.13 | 1.64 | -1.47 | 0.01 | 1.46 |
| 122 | 18 | 10.99<sub>2,15</sub>** | 1.76 | 19.39*** | -1.16 | -3.10 | 4.26 |
| 147 | 20 | 10.94<sub>2,17</sub>*** | 23.07*** | 1.57 | 2.14 | -3.47 | 1.33 |
| 271 | 19 | 4.01<sub>2,16</sub>* | 4.38‡ | 1.17 | -2.21 | 0.65 | 1.56 |
| 297 | 17 | 9.15<sub>2,14</sub>** | 8.25* | 2.48 | -4.74 | 0.86 | 3.88 |

Notes: \*\*\* $p < .001$ ; \*\* $p < .01$ ; \* $p < .05$ ; ‡ $p < .10$ . Sedentary behavior, SB; Physical activity, PA. Change, Δ. <sup>a</sup> Results only reported for participants with significant MANOVA results.

Supplemental Table 3 shows the MANOVA results across all participants, along with reconstructed differences in 24-hour movement behaviors on drinking versus non-drinking days. Across the 51 participants, 21 (41.2%) increased their sleep on drinking days, whereas 30 (58.8%) showed decreases in sleep on drinking days. Among participants who increased sleep on drinking days, sleep duration increased by an average of 0.80 hours, accompanied by reductions in SB (−0.72 hours) and PA (−0.07 hours). In contrast, participants who decreased sleep on drinking days slept 1.04 fewer hours, with time primarily reallocated to PA (+1.00 hours) and only minimal changes in SB.

## Discussion

This study examined day-to-day associations between alcohol use and device-based measures of sleep, as well as compositional changes across the 24-hour movement behaviors (sleep, SB, PA) on drinking versus non-drinking days. We observed a small dose–response pattern between alcohol use and sleep, with each additional alcoholic drink above the participant’s usual amount corresponding to approximately four fewer minutes of sleep. Additionally, participants slept 16 fewer minutes on drinking days compared to non-drinking days. There was substantial between-person variability in patterns of 24-hour movement behaviors across drinking versus non-drinking days, with 59% of participants reallocating time from sleep to PA on drinking days (1.00 more hours of PA), and 41% of participants reallocating time from SB to sleep on drinking days (0.72 fewer hours of SB).

While the dose-response findings in this study were relatively small, these findings are consistent with alcohol administration studies finding that acute alcohol consumption worsens sleep quantity in a dose-dependent manner [8–11]. When considering prior naturalistic research, the dose-response and drinking day findings aligned with one study finding that students slept less on binge drinking days compared to non-binge drinking days, though it should be noted that non-binge drinking days included both non-drinking days and days with alcohol use below the binge-drinking threshold [35]. However, our findings contrasted with other naturalistic studies finding no within-person associations between the amount of daily alcohol use and sleep duration [31–34,36,37]. Despite finding no effects on total sleep quantity, other naturalistic studies did find dose-response effects of alcohol use with other sleep characteristics, such as delayed bed and wake times, greater wakefulness after sleep onset, and worse sleep quality [31–36]. This study did not investigate sleep quality; however, given the decreased sleep quantity with greater alcohol consumption, our findings reinforce concerns about the acute detrimental effects of alcohol use on sleep.

Another notable finding in the current study was the between-person variability in alcohol use-sleep associations, with individual MANOVAs indicating that four participants showed significant differences in time use reallocation across drinking versus non-drinking days. This heterogeneity helps explain the non-significant between-person effects in our multi-level models, as well as conflicting findings from previous studies examining the between-person associations of alcohol use and sleep, with studies finding greater drinking corresponded with less average sleep time [32], greater average sleep time [33,36,37], or was not associated with average sleep time [34,35]. It underscores that, because different people respond differently, focusing on individuals rather than the entire populations can provide better insights into potential health impacts and behavioral changes. Unfortunately, drawing inferences from these previous studies is somewhat complicated due to the failure to account for the other 24-hour movement behaviors (PA, SB), both of which impact sleep quantity and quality [12–14].

A novel contribution of the current study was using wearable sensors to measure all 24-hour movement behaviors (sleep, SB, PA) and using CoDA to provide further insight into the between-person heterogeneity of how drinking days alter both sleep and the overall balance among daily movement behaviors. Although the sample size was modest, the MANOVAs and ternary plots revealed distinct within-person behavioral reallocations among five of the participants. Specifically, four participants decreased sleep time on drinking days, with time use generally reallocated to increased PA time, while one individual increased sleep time on drinking days, with decreased SB time. One plausible explanation for these patterns relates to the social and physical context of alcohol use. Alcohol use most commonly occurs in social settings, with population and daily diary studies indicating that over two-thirds of drinking occasions occur in a social context [53] and that solitary drinking occurs on less than 14% of days [54,55]. When considering physical context of alcohol use, daily diary studies indicate that over 50% of drinking occasions occur at bars, parties, or events [54]. Although no empirical data currently characterize movement behaviors during social drinking occasions specifically, these settings likely involve extended periods of standing, ambulation, and intermittent movement (e.g., circulating or socializing), behaviors that would typically be classified as LPA. Given that our PA variable encompassed light, moderate, and vigorous movement intensities, the observed reallocation of time from sleep to PA on drinking days may reflect, in part, increased accumulation of LPA related to the social and physical contexts of drinking occasions. It is also possible that these patterns reflect increases in MVPA, as some studies suggests that adults drinking more alcohol on days they engage in more leisure time PA [56], though the intensity of this activity may vary. Future studies incorporating device-based measures capable of distinguishing LPA from MVPA, alongside detailed contextual data about the social and physical environments of alcohol use, are needed to clarify whether these reallocations represent intentional engagement in leisure time PA, passive displacement into LPA, or a combination of both. Overall, these patterns suggest that the context of alcohol consumption may potentially impact broader shifts in 24-hour movement behaviors among some individuals, such that alcohol use occurring in more active social contexts (e.g., parties or events) may be associated with increased movement, whereas alcohol use in sedentary contexts (e.g., watching a movie) may not. Future studies investigating how social and environmental contexts of alcohol use shape 24-hour movement behaviors are warranted and could inform interventions, particularly given prior evidence that drinking intensity and the likelihood of engaging in protective behavioral strategies vary across contexts [57,58].

Although statistical significance was only achieved in five of the MANOVAs examining 24-hour time use on drinking versus non-drinking days, there were consistent trends observed, such that 59% of participants spent less time sleeping on drinking days, whereas 41% spent more time sleeping on drinking days. We believe that the lack of significant findings was primarily due to the limited analytic sample (n = 51) and modest variability in daily alcohol exposure. Future studies with larger samples and longer monitoring periods are warranted to clarify whether alcohol-induced sleep changes consistently reallocate time among SB and PA or whether individual differences—such as chronotype or drinking context—moderate these effects. Integrating wearable physiological data (e.g., heart rate variability or skin temperature) could also help elucidate mechanistic pathways linking alcohol metabolism and sleep fragmentation [59]. Combining contextual information with wearable measures also has the potential to inform interventions that are personalized to individuals’ needs and behavior patterns via N-of-1 trials or just-in-time adaptive interventions, which have become increasingly prevalent in precision and behavioral medicine over the past decade due to between-person heterogeneity in treatment responses [60–63].

### Strengths and limitations

A major strength of this study is the use of wearable devices to measure 24-hour movement behaviors collected over multiple days, combined with daily self-reports of alcohol use and sophisticated modelling of 24-hour movement behavior compositions (CoDA). This approach allows for within-person inference (day-level variation) and between-person differences, and addresses the interdependence of sleep, SB, and PA [12–15]. The recruitment of overweight/obese adults and the inclusion of light, moderate, and heavy alcohol users enhances the relevance of findings for public health and behavioral interventions.

There were several limitations to this study. First, the sample was limited to individuals with overweight/obesity, who may exhibit different 24-hour movement behavior compositions compared to individuals with normal weight [64], thereby limiting generalizability, and these associations may differ across BMI groups. Second, our analytic subsample for CoDA (n = 91 for multilevel models and n = 57 for MANOVAs) was relatively small and selected based on strict wear-time and drinking criteria, which may limit generalizability and reduce statistical power to detect compositional effects. Third, alcohol use was self-reported via daily morning surveys and may be subject to recall bias or under-reporting, though daily morning reports are a commonly used measure that strongly correlate with sensor-based measures of alcohol use [65]. Fourth, sleep was measured using the activPAL monitor, which overestimates sleep duration compared to self-report and other wearable sensors [66–69], which could explain differences between our findings and previous studies. However, the ActiWatch was discontinued prior to data collection for this study and the activPAL was selected due to its ability to provide reasonable estimates of all 24-hour movement behaviors [45,46]. Fifth, while our models adjusted for several covariates, other potential confounders (e.g., caffeine, other drug use, chronotype, sleep disorders) were not included in the models. Finally, the observational design precludes causal inference, and the sample was relatively homogeneous (region, age, overweight/obese adults), limiting broader generalizability.

### Conclusions

Our findings extend prior work by combining wearable measures of 24-hour movement behaviors with daily reports of alcohol use and applying compositional modeling to capture the daily associations between alcohol use and sleep while accounting for the interdependent nature of daily movement behaviors. The findings suggest that drinking days and higher-than-usual alcohol consumption may be associated with modest reductions in nightly sleep duration and subtly shift the balance of movement behaviors across the 24-hour day. Understanding these behavioral trade-offs may inform individualized behavioral interventions aimed at mitigating alcohol-related sleep disruption and promoting healthier daily movement patterns.

## Data Availability

The minimal data set and accompanying R code are available at Open Science Framework via https://osf.io/gfw73

## Acknowledgements

Thank you to members of the Health, Exercise, and Lifestyle (HEAL) Lab for helping with data collection. Thank you to all the individuals who participated in this study. Your participation was fundamental to advancing our research and knowledge.

## References

1. Hirshkowitz M, Whiton K, Albert SM, Alessi C, Bruni O, DonCarlos L, et al. National Sleep Foundation’s sleep time duration recommendations: methodology and results summary. Sleep Health. 2015;1:40–3. 10.1016/j.sleh.2014.12.010

2. Adams RJ, Appleton SL, Taylor AW, Gill TK, Lang C, McEvoy RD, et al. Sleep health of Australian adults in 2016: results of the 2016 Sleep Health Foundation national survey. Sleep Health. 2017;3:35–42. 10.1016/j.sleh.2016.11.005

3. Krueger PM, Friedman EM. Sleep Duration in the United States: A Cross-sectional Population-based Study. American Journal of Epidemiology. 2009;169:1052–63. 10.1093/aje/kwp023

4. Scott H, Naik G, Lechat B, Manners J, Fitton J, Nguyen DP, et al. Are we getting enough sleep? Frequent irregular sleep found in an analysis of over 11 million nights of objective in-home sleep data. Sleep Health. 2024;10:91–7. 10.1016/j.sleh.2023.10.016

5. Johnson EO, Roehrs T, Roth T, Breslau N. Epidemiology Of Alcohol and Medication As Aids To Sleep in Early Adulthood. Sleep. 1998;21:178–86. 10.1093/sleep/21.2.178

6. Roehrs T, Roth T. Sleep, sleepiness, and alcohol use. Alcohol Res Health. 2001;25:101–9.

7. Roehrs T, Roth T. Insomnia pharmacotherapy. Neurotherapeutics. 2012;9:728–38. 10.1007/s13311-012-0148-3

8. Colrain IM, Nicholas CL, Baker FC. Alcohol and the sleeping brain. Handb Clin Neurol. 2014;125:415–31. 10.1016/B978-0-444-62619-6.00024-0

9. Ebrahim IO, Shapiro CM, Williams AJ, Fenwick PB. Alcohol and Sleep I: Effects on Normal Sleep. Alcoholism Clin & Exp Res [Internet]. 2013 [cited 2025 Nov 20];37. 10.1111/acer.12006

10. Gardiner C, Weakley J, Burke LM, Roach GD, Sargent C, Maniar N, et al. The effect of alcohol on subsequent sleep in healthy adults: A systematic review and meta-analysis. Sleep Medicine Reviews. 2025;80:102030. 10.1016/j.smrv.2024.102030

11. Thakkar MM, Sharma R, Sahota P. Alcohol disrupts sleep homeostasis. Alcohol. 2015;49:299–310. 10.1016/j.alcohol.2014.07.019

12. Dumuid D, Stanford TE, Martin-Fernández J-A, Pedišiæ Ž, Maher CA, Lewis LK, et al. Compositional data analysis for physical activity, sedentary time and sleep research. Stat Methods Med Res. 2018;27:3726–38. 10.1177/0962280217710835

13. Dumuid D, Pedišiæ Ž, Palarea-Albaladejo J, Martín-Fernández JA, Hron K, Olds T. Compositional Data Analysis in Time-Use Epidemiology: What, Why, How. IJERPH. 2020;17:2220. 10.3390/ijerph17072220

14. Matricciani L, Bin YS, Lallukka T, Kronholm E, Wake M, Paquet C, et al. Rethinking the sleep-health link. Sleep Health. 2018;4:339–48. 10.1016/j.sleh.2018.05.004

15. Pedišiæ Ž. Measurement issues and poor adjustments for physical activity and sleep undermine sedentary behaviour research—the focus should shift to the balance between sleep, sedentary behaviour, standing and activity. Kinesiology. 2014;46:135–46.

16. Kredlow MA, Capozzoli MC, Hearon BA, Calkins AW, Otto MW. The effects of physical activity on sleep: a meta-analytic review. J Behav Med. 2015;38:427–49. 10.1007/s10865-015-9617-6

17. Xie Y, Liu S, Chen X-J, Yu H-H, Yang Y, Wang W. Effects of Exercise on Sleep Quality and Insomnia in Adults: A Systematic Review and Meta-Analysis of Randomized Controlled Trials. Front Psychiatry. 2021;12:664499. 10.3389/fpsyt.2021.664499

18. Duquet L, Galli S, Haffen E, Giustiniani J. The Impact of Physical Activity on Sleep in Alcohol Users: A Systematic Review. Addiction Biology. 2025;30:e70050. 10.1111/adb.70050

19. Ju K, Liu N, Wu R, Shi X. The relationship between sedentary behavior, sleep duration, and sleep disorders: analysis of the 2007–2014 National Health and Nutrition Examination Survey. Front Neurol. 2025;16:1488443. 10.3389/fneur.2025.1488443

20. Yang Y, Shin JC, Li D, An R. Sedentary Behavior and Sleep Problems: a Systematic Review and Meta-Analysis. IntJ Behav Med. 2017;24:481–92. 10.1007/s12529-016-9609-0

21. Brown DMY, Burkart S, Groves CI, Balbim GM, Pfledderer CD, Porter CD, et al. A systematic review of research reporting practices in observational studies examining associations between 24-h movement behaviors and indicators of health using compositional data analysis. JASSB. 2024;3:23. 10.1186/s44167-024-00062-8

22. Chastin SFM, Palarea-Albaladejo J, Dontje ML, Skelton DA. Combined Effects of Time Spent in Physical Activity, Sedentary Behaviors and Sleep on Obesity and Cardio-Metabolic Health Markers: A Novel Compositional Data Analysis Approach. Devaney J, editor. PLoS ONE. 2015;10:e0139984. 10.1371/journal.pone.0139984

23. Evenson KR, Goto MM, Furberg RD. Systematic review of the validity and reliability of consumer-wearable activity trackers. Int J Behav Nutr Phys Act. 2015;12:159. 10.1186/s12966-015-0314-1

24. Giurgiu M, Timm I, Becker M, Schmidt S, Wunsch K, Nissen R, et al. Quality Evaluation of Free-living Validation Studies for the Assessment of 24-Hour Physical Behavior in Adults via Wearables: Systematic Review. JMIR Mhealth Uhealth. 2022;10:e36377. 10.2196/36377

25. Lee T, Cho Y, Cha KS, Jung J, Cho J, Kim H, et al. Accuracy of 11 Wearable, Nearable, and Airable Consumer Sleep Trackers: Prospective Multicenter Validation Study. JMIR Mhealth Uhealth. 2023;11:e50983. 10.2196/50983

26. Rosenberger ME, Buman MP, Haskell WL, Mcconnell MV, Carstensen LL. Twenty-four hours of sleep, sedentary behavior, and physical activity with nine wearable devices. Medicine & Science in Sports & Exercise. 2016;48:457–65. 10.1249/MSS.0000000000000778

27. Scott H, Lack L, Lovato N. A systematic review of the accuracy of sleep wearable devices for estimating sleep onset. Sleep Medicine Reviews. 2020;49:101227. 10.1016/j.smrv.2019.101227

28. Van De Water ATM, Holmes A, Hurley DA. Objective measurements of sleep for non-laboratory settings as alternatives to polysomnography - a systematic review: A systematic review of objective sleep measures. Journal of Sleep Research. 2011;20:183–200. 10.1111/j.1365-2869.2009.00814.x

29. Bolger N, Laurenceau J-P. Intensive longitudinal methods: An introduction to diary and experience sampling research. New York: Guilford Press; 2013.

30. Collins LM. Analysis of Longitudinal Data: The Integration of Theoretical Model, Temporal Design, and Statistical Model. Annu Rev Psychol. 2006;57:505–28. 10.1146/annurev.psych.57.102904.190146

31. De Rosa O, Menghini L, Kerr E, Müller-Oehring E, Nooner K, Hasler BP, et al. Exploring the relationship between sleep patterns, alcohol and other substances consumption in young adults: Insights from wearables and Mobile surveys in the National Consortium on alcohol and NeuroDevelopment in adolescence (NCANDA) cohort. International Journal of Psychophysiology. 2025;209:112524. 10.1016/j.ijpsycho.2025.112524

32. Fucito LM, Bold KW, Van Reen E, Redeker NS, O’Malley SS, Hanrahan TH, et al. Reciprocal variations in sleep and drinking over time among heavy-drinking young adults. Journal of Abnormal Psychology. 2018;127:92–103. 10.1037/abn0000312

33. Lydon DM, Ram N, Conroy DE, Pincus AL, Geier CF, Maggs JL. The within-person association between alcohol use and sleep duration and quality in situ: An experience sampling study. Addictive Behaviors. 2016;61:68–73. 10.1016/j.addbeh.2016.05.018

34. Miller MB, Freeman LK, Deroche CB, Park CJ, Hall NA, McCrae CS. Sleep and alcohol use among young adult drinkers with Insomnia: A daily process model. Addictive Behaviors. 2021;119:106911. 10.1016/j.addbeh.2021.106911

35. Patrick ME, Griffin J, Huntley ED, Maggs JL. Energy Drinks and Binge Drinking Predict College Students’ Sleep Quantity, Quality, and Tiredness. Behav Sleep Med. 2018;16:92–105. 10.1080/15402002.2016.1173554

36. Tracy EL, Reid KJ, Baron KG. The relationship between sleep and physical activity: the moderating role of daily alcohol consumption. Sleep. 2021;44:zsab112. 10.1093/sleep/zsab112

37. Tussey EJ, Wong MM. Bidirectional relationships between daily sleep and alcohol use. Alcohol. 2025;129:50–7. 10.1016/j.alcohol.2025.09.002

38. Lowder K, Courtney JB. Moderate-to-heavy habitual alcohol use appears to be detrimentally associated with inflammation even when accounting for diet quality in humans: findings from an observational intensive longitudinal study. Alcohol and Alcoholism. 2026;61:agag015. 10.1093/alcalc/agag015

39. Harris PA, Taylor R, Thielke R, Payne J, Gonzalez N, Conde JG. Research electronic data capture (REDCap)—A metadata-driven methodology and workflow process for providing translational research informatics support. Journal of Biomedical Informatics. 2009;42:377–81. 10.1016/j.jbi.2008.08.010

40. Garcia JB, Kerr ZY, Kyaw EM, Courtney JB. Investigating bidirectional within- and between-person associations between daily anxiety and physical activity participation. Psychology of Sport and Exercise. 2026;83:103027. 10.1016/j.psychsport.2025.103027

41. Hamilton CM, Strader LC, Pratt JG, Maiese D, Hendershot T, Kwok RK, et al. The PhenX Toolkit: Get the most from your measures. American Journal of Epidemiology. 2011;174:253–60. 10.1093/aje/kwr193

42. Collins RL, Parks GA, Marlatt GA. Social determinants of alcohol consumption: The effects of social interaction and model status on the self-administration of alcohol. J Consult Clin Psychol. 1985;53:189–200. 10.1037//0022-006x.53.2.189.%20PMID:%203998247

43. Piumatti G, Aresi G, Marta E. A psychometric analysis of the Daily Drinking Questionnaire in a nationally representative sample of young adults from a Mediterranean drinking culture. Journal of Ethnicity in Substance Abuse. 2023;22:171–88. 10.1080/15332640.2021.1918600

44. McKenna H, Treanor C, O’Reilly D, Donnelly M. Evaluation of the psychometric properties of self-reported measures of alcohol consumption: a COSMIN systematic review. Subst Abuse Treat Prev Policy. 2018;13:6. 10.1186/s13011-018-0143-8

45. Lyden K, Keadle SK, Staudenmayer J, Freedson PS. The activPALTM Accurately Classifies Activity Intensity Categories in Healthy Adults. Med Sci Sports Exerc. 2017;49:1022–8. 10.1249/MSS.0000000000001177

46. PAL Technologies Ltd – Providing the Evidence [Internet]. 2010 [cited 2023 Nov 29]. https://www.palt.com/. Accessed 29 Nov 2023

47. Hibbing PR, Carlson JA, Simon SL, Melanson EL, Creasy SA. Convergent Validity of Time in Bed Estimates From activPAL and Actiwatch in Free-Living Youth and Adults. Journal for the Measurement of Physical Behaviour. 2023;6:213–22. 10.1123/jmpb.2023-0011

48. Carlson JA, Tuz-Zahra F, Bellettiere J, Ridgers ND, Steel C, Bejarano C, et al. Validity of Two Awake Wear-Time Classification Algorithms for activPAL in Youth, Adults, and Older Adults. J Meas Phys Behav. 2021;4:151–62. 10.1123/jmpb.2020-0045

49. Marshall S, Nicaise V, Ji M, Huerta C, Haubenstricker J, Levy S, et al. Using step cadence goals to increase moderate-to-vigorous-intensity physical activity. Medicine and science in sports and exercise [Internet]. Med Sci Sports Exerc; 2013 [cited 2023 Nov 27];45. 10.1249/MSS.0b013e318277a586

50. Tudor-Locke C, Aguiar EJ, Han H, Ducharme SW, Schuna JM, Barreira TV, et al. Walking cadence (steps/min) and intensity in 21–40 year olds: CADENCE-adults. Int J Behav Nutr Phys Act. 2019;16:8. 10.1186/s12966-019-0769-6

51. Aitchison J. Principles of compositional data analysis. IMS Lecture Notes - Monograph Series. 1994;24:73–81.

52. Aitchison J. A concise guide to compositional data analysis [Internet]. 2005. https://eprints.gla.ac.uk/259608/

53. Creswell KG, Fairbairn CE. The need for multi-participant alcohol administration studies. Addiction. 2025;120:574–7. 10.1111/add.16735

54. Terry-McElrath YM, Arterberry BJ, Patrick ME. Alcohol use contexts (social settings, drinking games/specials, and locations) as predictors of high-intensity drinking on a given day among U.S. young adults. Alcohol: Clinical and Experimental Research. 2023;47:273–84. 10.1111/acer.14985

55. Ally AK, Lovatt M, Meier PS, Brennan A, Holmes J. Developing a social practice-based typology of British drinking culture in 2009-2011: implications for alcohol policy analysis: Typology of British drinking practices. Addiction. 2016;111:1568–79. 10.1111/add.13397

56. Conroy DE, Ram N, Pincus AL, Coffman DL, Lorek AE, Rebar AL, et al. Daily physical activity and alcohol use across the adult lifespan. Health Psychol. 2015;34:653–60. 10.1037/hea0000157.%20PMID:%2025222084;%20PMCID:%20PMC4362843

57. Cox MJ, Stevens AK, Janssen T, Jackson KM. Event-level contextual predictors of high-intensity drinking events among young adults. Drug and Alcohol Dependence. 2022;239:109590. 10.1016/j.drugalcdep.2022.109590

58. Cox MJ, Johnson L, Roudebush M, Godbole A, Egan KL. Likelihood of Young Adult Engagement in Protective Behavioral Strategies for Alcohol Use across Drinking Contexts: Implications for Adaptive Interventions. Substance Use & Misuse. 2024;59:902–9. 10.1080/10826084.2024.2310484

59. Strüven A, Schlichtiger J, Hoppe JM, Thiessen I, Brunner S, Stremmel C. The Impact of Alcohol on Sleep Physiology: A Prospective Observational Study on Nocturnal Resting Heart Rate Using Smartwatch Technology. Nutrients. 2025;17:1470. 10.3390/nu17091470

60. Augustine EF, Yu TW, Finkel RS. N-of-1 Studies in an Era of Precision Medicine. JAMA. 2024;332:1386. 10.1001/jama.2024.14637

61. Hsu TC, Whelan P, Gandrup J, Armitage CJ, Cordingley L, McBeth J. Personalized interventions for behaviour change: A scoping review of just-in-time adaptive interventions. British J Health Psychol. 2025;30:e12766. 10.1111/bjhp.12766

62. Lillie EO, Patay B, Diamant J, Issell B, Topol EJ, Schork NJ. The n-of-1 clinical trial: the ultimate strategy for individualizing medicine? Per Med. 2011;8:161–73. 10.2217/pme.11.7

63. Perski O, Hébert ET, Naughton F, Hekler EB, Brown J, Businelle MS. Technology-mediated just-in-time adaptive interventions (JITAIs) to reduce harmful substance use: a systematic review. Addiction. 2022;117:1220–41. 10.1111/add.15687

64. Chastin SFM, Palarea-Albaladejo J, Dontje ML, Skelton DA. Combined Effects of Time Spent in Physical Activity, Sedentary Behaviors and Sleep on Obesity and Cardio-Metabolic Health Markers: A Novel Compositional Data Analysis Approach. Devaney J, editor. PLoS ONE. 2015;10:e0139984. 10.1371/journal.pone.0139984

65. Russell MA, Turrisi RJ, Smyth JM. Transdermal sensor features correlate with ecological momentary assessment drinking reports and predict alcohol-related consequences in young adults’ natural settings. Alcohol Clin & Exp Res. 2022;46:100–13. 10.1111/acer.14739

66. Courtney JB, Nuss K, Lyden K, Harrall KK, Glueck DH, Villalobos A, et al. Comparing the activPAL Software’s Primary Time in Bed Algorithm against Self-Report and van Der Berg’s Algorithm. Measurement in Physical Education and Exercise Science. 2021;25:212–26. 10.1080/1091367X.2020.1867146

67. Hibbing PR, Carlson JA, Simon SL, Melanson EL, Creasy SA. Convergent validity of time in bed estimates from activPAL and Actiwatch in free-living youth and adults. J Meas Phys Behav. 2023;6:213–22. 10.1123/jmpb.2023-0011

68. Inan-Eroglu E, Powell L, Hamer M, O’Donovan G, Duncan MJ, Stamatakis E. Is There a Link between Different Types of Alcoholic Drinks and Obesity? An Analysis of 280,183 UK Biobank Participants. IJERPH. 2020;17:5178. 10.3390/ijerph17145178

69. Leister KR, Garay J, Barreira TV. Validity of a Novel Algorithm to Detect Bedtime, Wake Time, and Sleep Time in Adults. Journal for the Measurement of Physical Behaviour. 2022;5:76–84. 10.1123/jmpb.2021-0027

